# Patient Attitudes About Light Therapy and Negative Ion Therapy for Nonseasonal Depression: An Online Survey Study

**DOI:** 10.1101/2021.09.29.21264324

**Authors:** Iman Lahouaoula, Victor W. Li, Aidan Scott, André Do, Erin E. Michalak, Jill K. Murphy, Samantha Huang, Raymond W. Lam

## Abstract

**Objective:** No studies have yet evaluated whether light therapy or negative ion therapy can be used as maintenance treatment after acute treatment with antidepressants in patients with major depressive disorder. To address the importance of this question, we surveyed participants with depression to determine their knowledge and attitudes about light therapy and negative ion therapy, and their willingness to participate in a randomized clinical trial with these therapies substituting for antidepressants for maintenance treatment.

**Methods:** Participants with a self-reported diagnosis of depression were recruited by email, newsletters, and social media to complete an online survey with questions about awareness and effectiveness of light therapy and negative ion therapy for depression. Vignettes describing the use of these therapies for maintenance treatment were presented with follow up questions about the ease of use and reasons for wanting (and not wanting) to use the therapies instead of antidepressants. Another vignette described a randomized study with these therapies followed by questions on whether participants would likely volunteer for the study. Chi-square tests were used to examine differences in responses between therapies.

**Results:** A total of 221 participants completed the survey. Most of them were aware of both therapies, but more participants had heard of light therapy (95% compared to 62% for negative ion therapy, p<0.0001), had used light therapy (28% versus 16%, p<0.003), and regarded light therapy as effective (54% versus 37%, p<0.001). Both therapies were considered easy to use. The majority of participants (78%) thought that it was important to find non-medication therapies for maintenance treatment, and 77% responded that they would likely volunteer for a randomized study to determine efficacy of the two therapies for maintenance treatment.

**Conclusion:** People with depression are generally aware of light therapy and negative ion therapy and believe they would be good therapies to substitute for antidepressants in maintenance treatment. These findings support the importance and feasibility for a randomized relapse prevention trial with light therapy and negative ion therapy in patients with depression.

## INTRODUCTION

Major depressive disorder (MDD) affects approximately 264 million people worldwide and is one of the top 3 medical causes of years lived with disability (GBD 2017 Collaborators, 2018). Although there are many evidence-based treatment options for MDD, antidepressants are recommended first-line treatments for moderate to severe depression (Kennedy et al, 2016). Once patients are recovered from the acute episode, maintenance antidepressant treatment is recommended for 6-24 months, or longer, to prevent relapse and recurrence (Kennedy et al, 2016). Several meta-analyses of randomized controlled trials (RCTs) have confirmed that maintenance antidepressants are effective to prevent relapse (Sim et al, 2016; Kato et al, 2021).

Despite the evidence for maintenance, however, patients often discontinue antidepressants too early after acute treatment, putting them at risk of relapse and consequent impairment in functioning and quality of life (Olfson et al, 2006). Some studies suggest that less than 30% of patients continue antidepressants for more than three months (Li et al, 2016). Reasons for stopping antidepressants too soon include persistent side effects such as sexual dysfunction and weight gain, cost, adverse effects with long term use of antidepressants (e.g., osteoporosis, gastrointestinal bleeding, risk of drug interactions with medications for other medical conditions), and patient preference for non-medication treatments (Samples and Mojtabai, 2015).

Given the reluctance for longer-term use of antidepressants, perhaps patients would consider using non-medication treatments to substitute for antidepressants during maintenance treatment. Non-medication treatments that have been studied for depression include bright light therapy and negative ion therapy. Light therapy has long been considered a first-line treatment for seasonal affective disorder (SAD), but increasing evidence supports its efficacy in nonseasonal MDD (Lam et al, 2016; Tao et al., 2020). Studies of high-density negative ion treatment have also shown positive results in reducing depressive symptoms (Goel et al., 2005; Perez et al, 2013). Both these treatments are safe and well-tolerated, with relatively mild side effects (Terman & Terman, 2006; Goel et al., 2005).

Light therapy and negative ion therapy may be candidates for antidepressant substitution, but, given that neither treatment has been studied for relapse prevention, rigorous RCTs are needed to demonstrate efficacy and safety. However, it is unclear whether patients would find it acceptable to use one of these treatments while discontinuing antidepressants, especially since these treatments are not widely used in clinical practice. Hence, we surveyed people with depression to determine (1) the importance of the clinical question from a patient perspective, (2) their familiarity with light and ion therapy, and (3) the feasibility of conducting an RCT with light and ion therapy for maintenance treatment of MDD.

## METHODS

### Participants

This online survey study received approval from the Behavioural Research Ethics Board at the University of British Columbia. Participants were recruited from various sources, including social media postings, newsletters and research networks in Canada. Inclusion criteria were: (1) age 19-65 years; (2) diagnosed with depression by physician or psychologist, by self-report; (3) capable of informed consent; (4) access to an internet-enabled computer or mobile device; (5) able to read and understand English.

### Survey Methods

All survey data was collected electronically using Qualtrics. Participants provided informed consent before starting the online survey. The survey consisted of 3 sections. Section 1 included questions about demographics. Section 2 addressed knowledge and attitudes about light and ion treatments. Information stems were first presented (see Appendix 1), such as “Light therapy is a treatment for depression that uses daily exposure to bright light from a light box device used at home. Light therapy usually has fewer side effects than antidepressant medications,” followed by questions “have you heard of light therapy for depression?” and “in your opinion, how effective is light therapy for depression treatment?” The latter question included responses: “More effective than antidepressants,” “As effective as antidepressants,” “Less effective than antidepressants,” “Not effective,” and “Unsure.” A similar stem and the same questions were presented for negative ion therapy.

Section 3 addressed substitution treatments for antidepressants in maintenance treatment of depression. Following a scenario for antidepressant maintenance treatment (Appendix 1), participants were asked, “In your opinion, how important is it to substitute an evidence-based <u>non-medication</u> treatment for antidepressants for maintenance treatment?” with responses of “Very important,” “Quite important,” “Somewhat important,” and “Not at all important.” Then, two vignettes with identical phrasing were presented for substituting light and ion therapy for maintenance treatment (Appendix 1), each followed by a question, “How easy would it be for you to use light/ion therapy as described?”. survey also included vignettes (Appendix 1) to illustrate an individual’s use of light/ion therapy, describing the cost, time frame and schedule, each followed by a question, “how easy would it be for you to use light/ion therapy as described?” with responses on a 7-point Likert scale ranging from “Very easy” to “Very difficult.” Further questions asked about reasons why they would want or not want the treatments. A final vignette was presented with the procedures for an RCT of active versus inactive treatment with light and ion therapy for maintenance treatment (Appendix 1), followed by a question asking, “How likely would you be to volunteer for this study?” with responses on a 7-point Likert scale ranging from “Very likely” to “Very unlikely.”

The data are reported in counts and percentages, not including missing data. Chi-square tests were used to compare frequencies, with significance level set at less than 0.05.

## RESULTS

A total of 221 individuals responded to the survey. Table 1 shows the demographic data of this sample. The majority of the total sample were female (68%), age 25-49 years (58%), self-identified as white/European (72%), had an undergraduate degree (46%), were employed full time (42%) and lived in an urban area (56%).

**Table 1.**
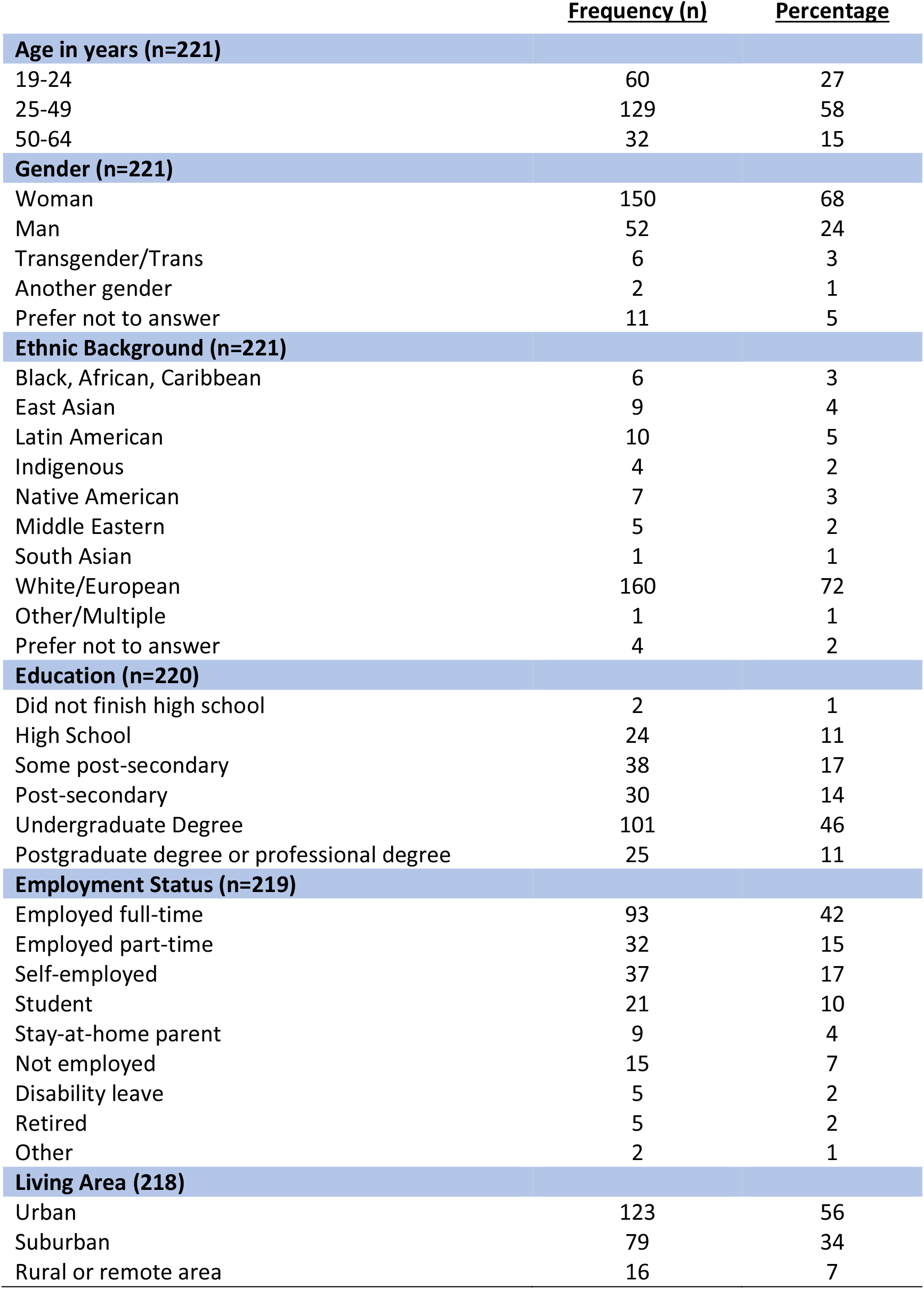
Demographic Characteristics. (number in brackets indicates responses)

|  | **Frequency (n)** | **Percentage** |
| --- | --- | --- |
| **Age in years (n=221)** |  |  |
| 19-24 | 60 | 27 |
| 25-49 | 129 | 58 |
| 50-64 | 32 | 15 |
| **Gender (n=221)** |  |  |
| Woman | 150 | 68 |
| Man | 52 | 24 |
| Transgender/Trans | 6 | 3 |
| Another gender | 2 | 1 |
| Prefer not to answer | 11 | 5 |
| **Ethnic Background (n=221)** |  |  |
| Black, African, Caribbean | 6 | 3 |
| East Asian | 9 | 4 |
| Latin American | 10 | 5 |
| Indigenous | 4 | 2 |
| Native American | 7 | 3 |
| Middle Eastern | 5 | 2 |
| South Asian | 1 | 1 |
| White/European | 160 | 72 |
| Other/Multiple | 1 | 1 |
| Prefer not to answer | 4 | 2 |
| **Education (n=220)** |  |  |
| Did not finish high school | 2 | 1 |
| High School | 24 | 11 |
| Some post-secondary | 38 | 17 |
| Post-secondary | 30 | 14 |
| Undergraduate Degree | 101 | 46 |
| Postgraduate degree or professional degree | 25 | 11 |
| **Employment Status (n=219)** |  |  |
| Employed full-time | 93 | 42 |
| Employed part-time | 32 | 15 |
| Self-employed | 37 | 17 |
| Student | 21 | 10 |
| Stay-at-home parent | 9 | 4 |
| Not employed | 15 | 7 |
| Disability leave | 5 | 2 |
| Retired | 5 | 2 |
| Other | 2 | 1 |
| **Living Area (218)** |  |  |
| Urban | 123 | 56 |
| Suburban | 79 | 34 |
| Rural or remote area | 16 | 7 |

In terms of treatment, 105 (48%) respondents had taken antidepressants in the past, 99 (45%) were currently taking antidepressants, and 17 (8%) had not taken antidepressants. Of those currently taking antidepressants (n=99), the length of time taking medication included less than 6 months (23%), 6-12 months (22%), and more than 24 months (46, 47%). Most respondents were satisfied with their antidepressant treatment (Very satisfied, 23%; Slightly satisfied, 43%; Neither satisfied nor dissatisfied, 22%; Slightly unsatisfied, 8%; Very unsatisfied, 2%). The majority also were knowledgeable of depression treatments in general, with 64 (29%) responding “Very knowledgeable,” 120 (54%) responding “Moderately knowledgeable,” and 25 (16%) responding “Somewhat knowledgeable.”

Most of the respondents (n=208, 95%) had heard of light therapy for depression, while fewer had heard of negative ion therapy (n=136, 62%) (χ=69.3, df=1, p<0.0001). Also, more participants had used light therapy (n=61, 28%) than had used negative ion therapy (n=35, 16%) (χ=8.9, df=1, p<0.003). Figure 1 shows the percentage responses for effectiveness of light therapy and negative ion therapy (data in Supplemental Table S1). The number of respondents that regarded light therapy as “more effective” or “as effective” as antidepressants was 119 (54%), while for negative ion therapy it was 81 (37%) (χ=13.3, df=1, p<0.001). There were also more respondents who were “Unsure” of effectiveness of negative ion therapy than light therapy (34% versus 19%, χ=12.8, df=1, p<0.001).

**Figure 1.**
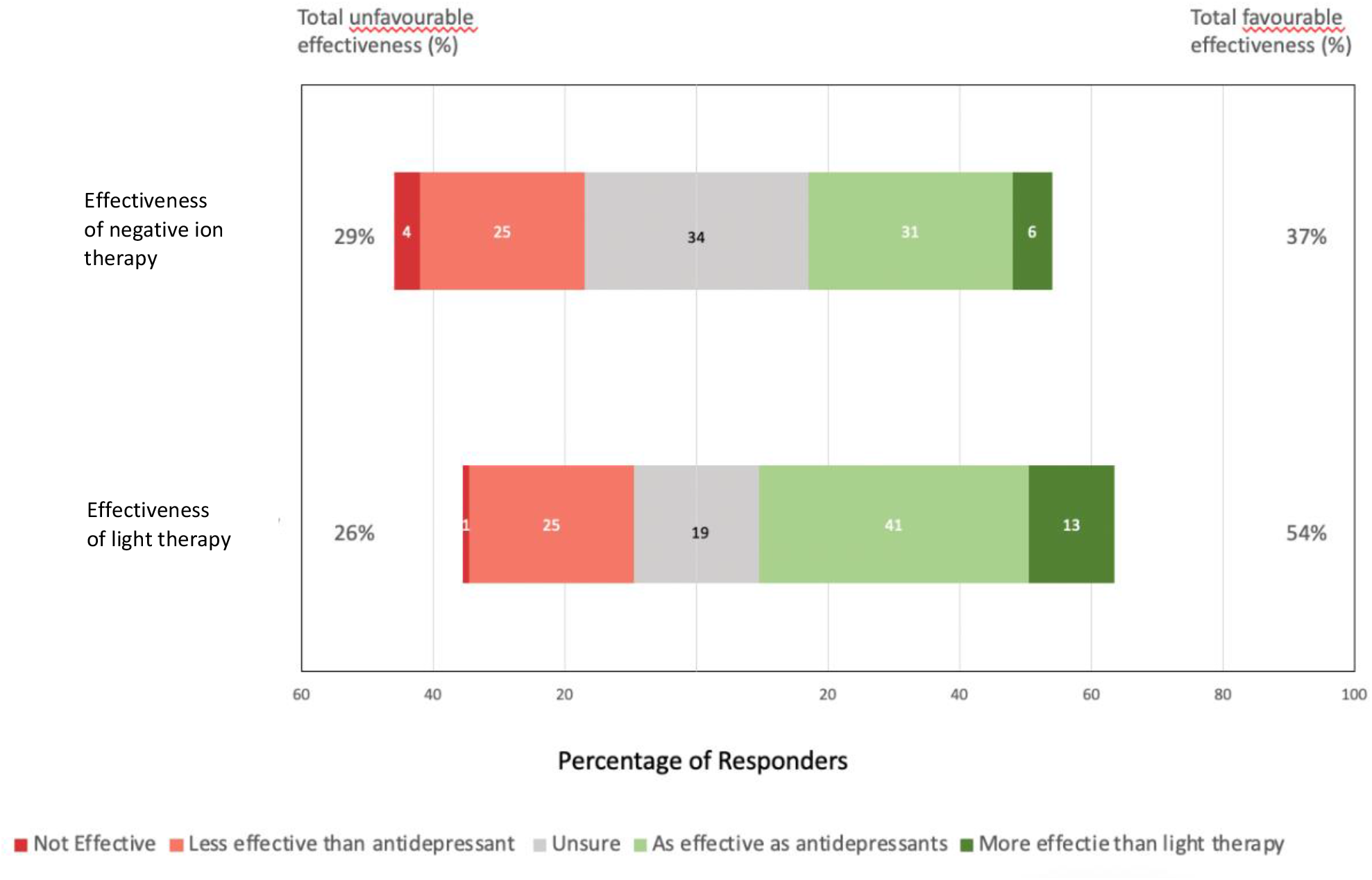
Responses on effectiveness of negative ion therapy and light therapy.

Figure 2 shows the responses for ease of use of the two therapies for maintenance treatment (data in Supplemental Table S1). Light therapy was regarded by 193 (88%) participants as slightly to very easy-to-use, while 176 (82%) thought negative ion therapy would be slightly to very easy-to-use (χ=3.3, df=1, p>0.05).

**Figure 2.**
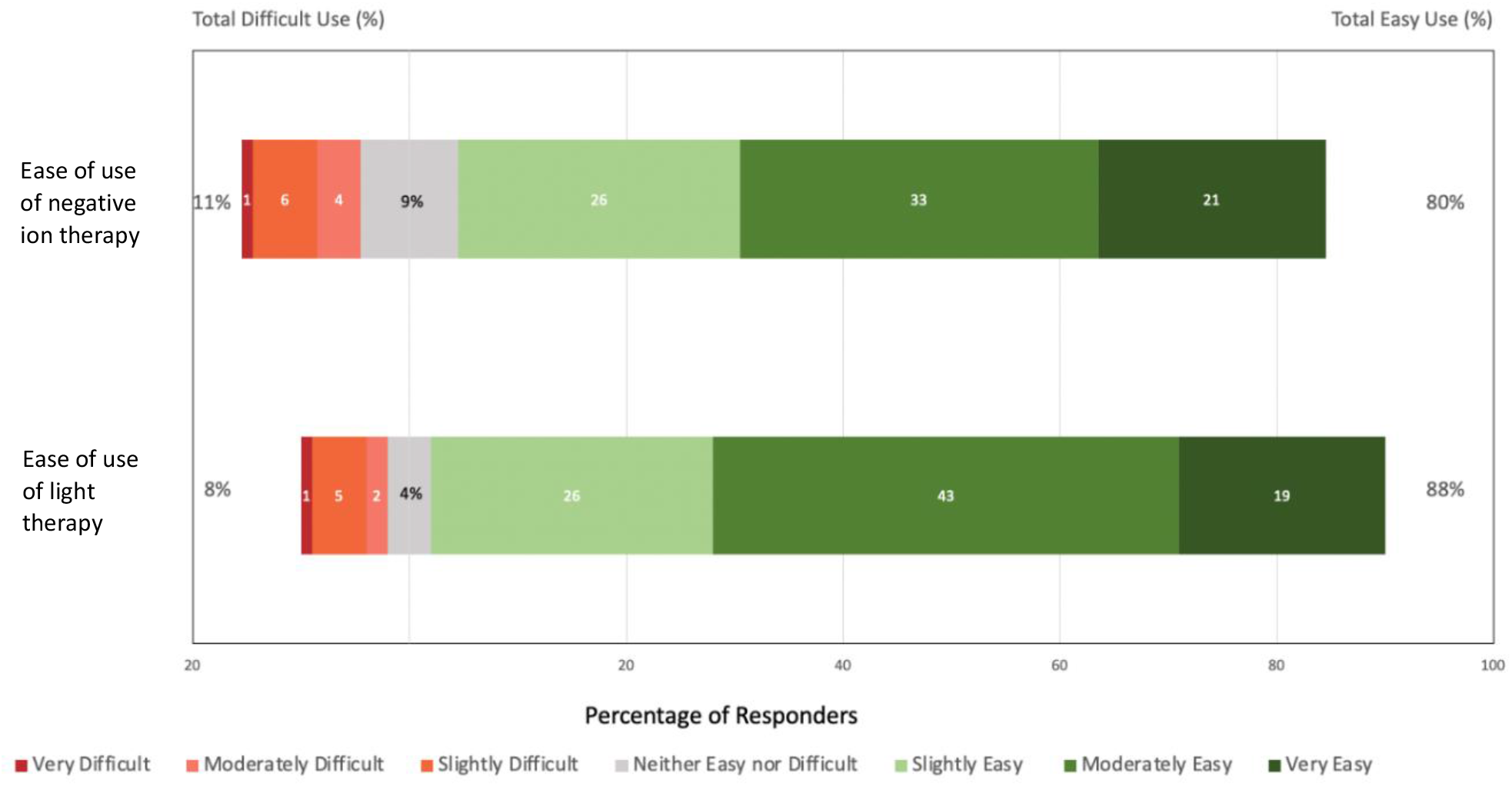
Responses about ease of use of negative ion therapy and light therapy.

Figures 3 and 4 show the main reasons for wanting to use and reasons for not wanting to use light therapy or negative ion therapy for maintenance treatment (data in Supplemental Table S1). The top three reasons for wanting use light or negative ion therapy instead of antidepressants were “it has fewer side effects,” “it is a non-medication treatment,” and “I don’t like taking medications. The top three reasons for not wanting to use light therapy or negative ion therapy were “I’m worried it won’t work,” “I’m worried about the withdrawal effects from stopping the antidepressants,” and “I don’t have time to do the therapy.”

**Figure 3.**
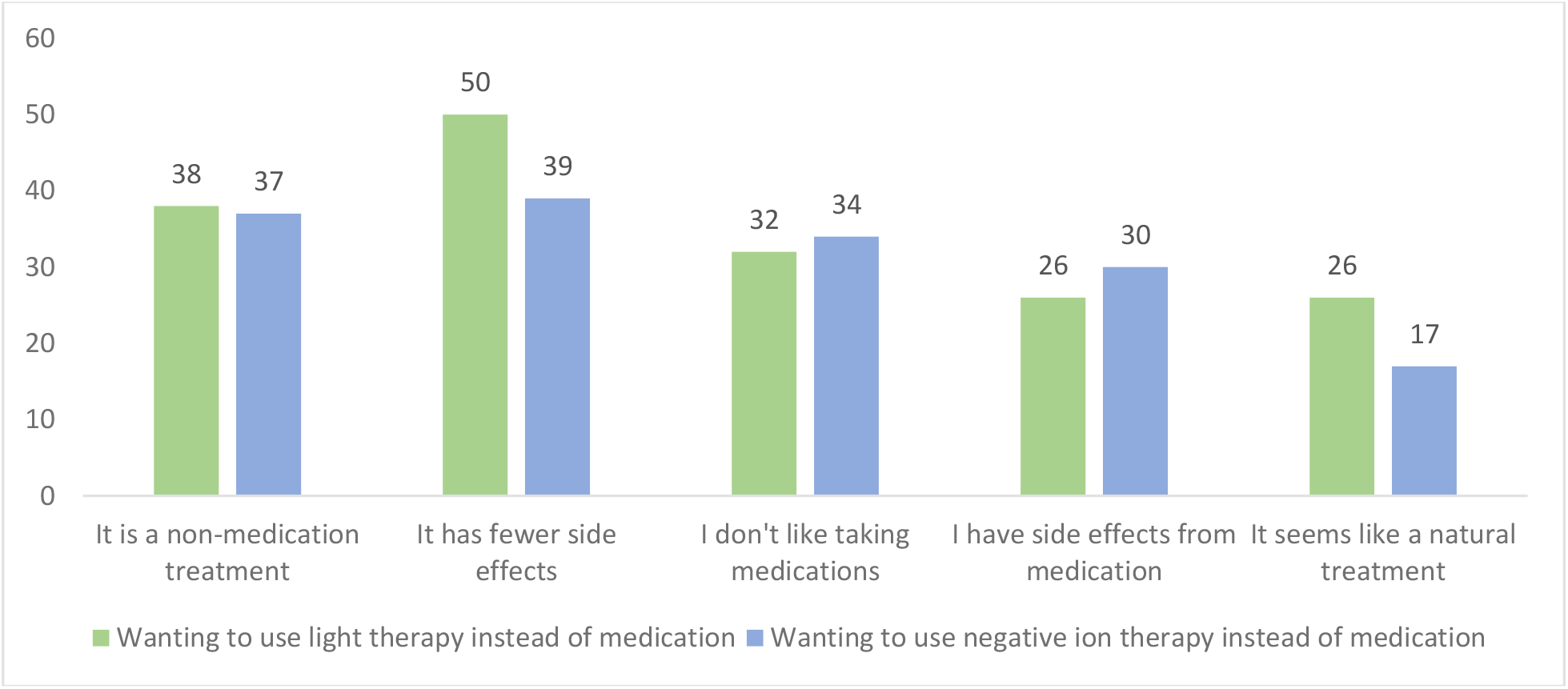
Reasons for wanting to use light therapy or negative ion therapy for maintenance treatment.

**Figure 4.**
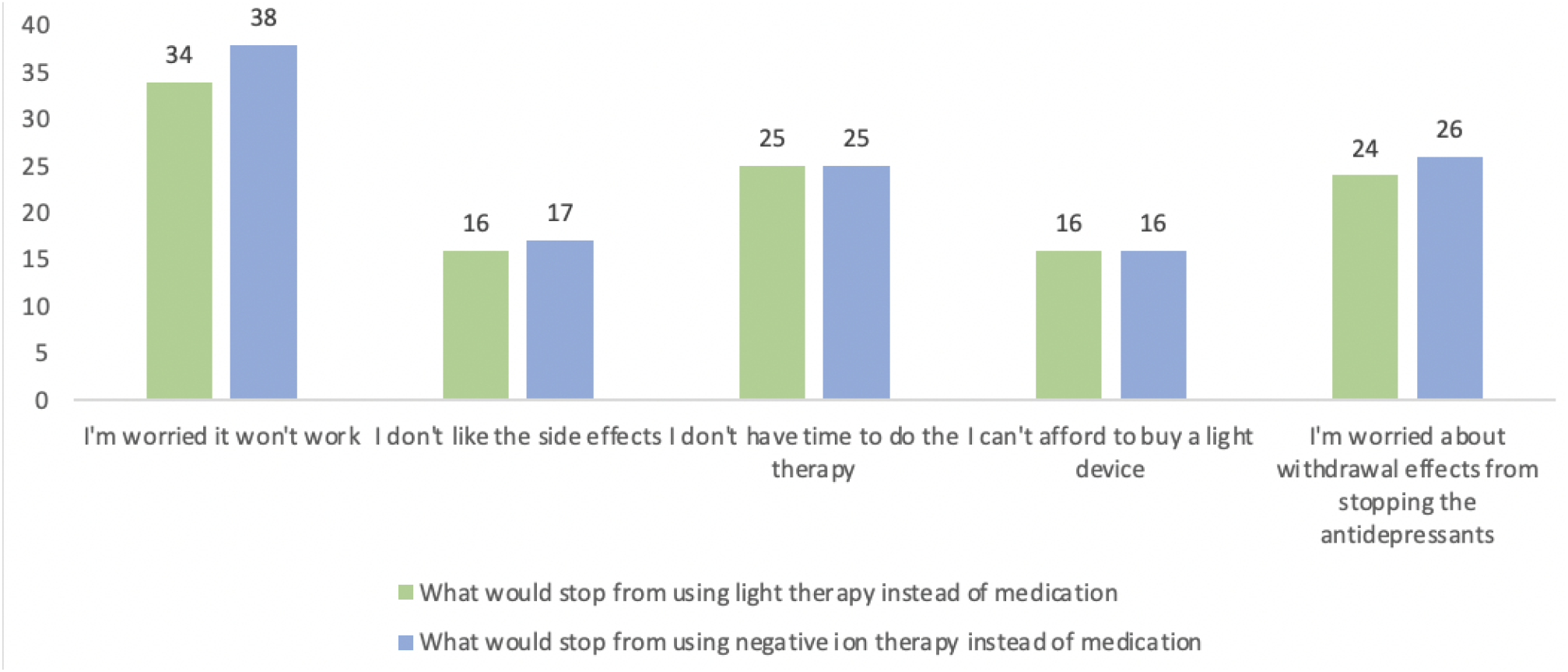
Reasons for not wanting to use light or negative ion therapies in maintenance treatment.

Figure 5 shows the responses about the importance of finding substitute non-medication treatments for maintenance treatment (data in Supplemental Table S2). Generally, support for the need for non-medication therapies was strong with 177 (78%) of participants identifying it as either “Very important” or “Quite important,” another 32 (15%) regarding it as “Somewhat important,” and only 14 (6%) viewing it as “Not at all important.”

**Figure 5.**
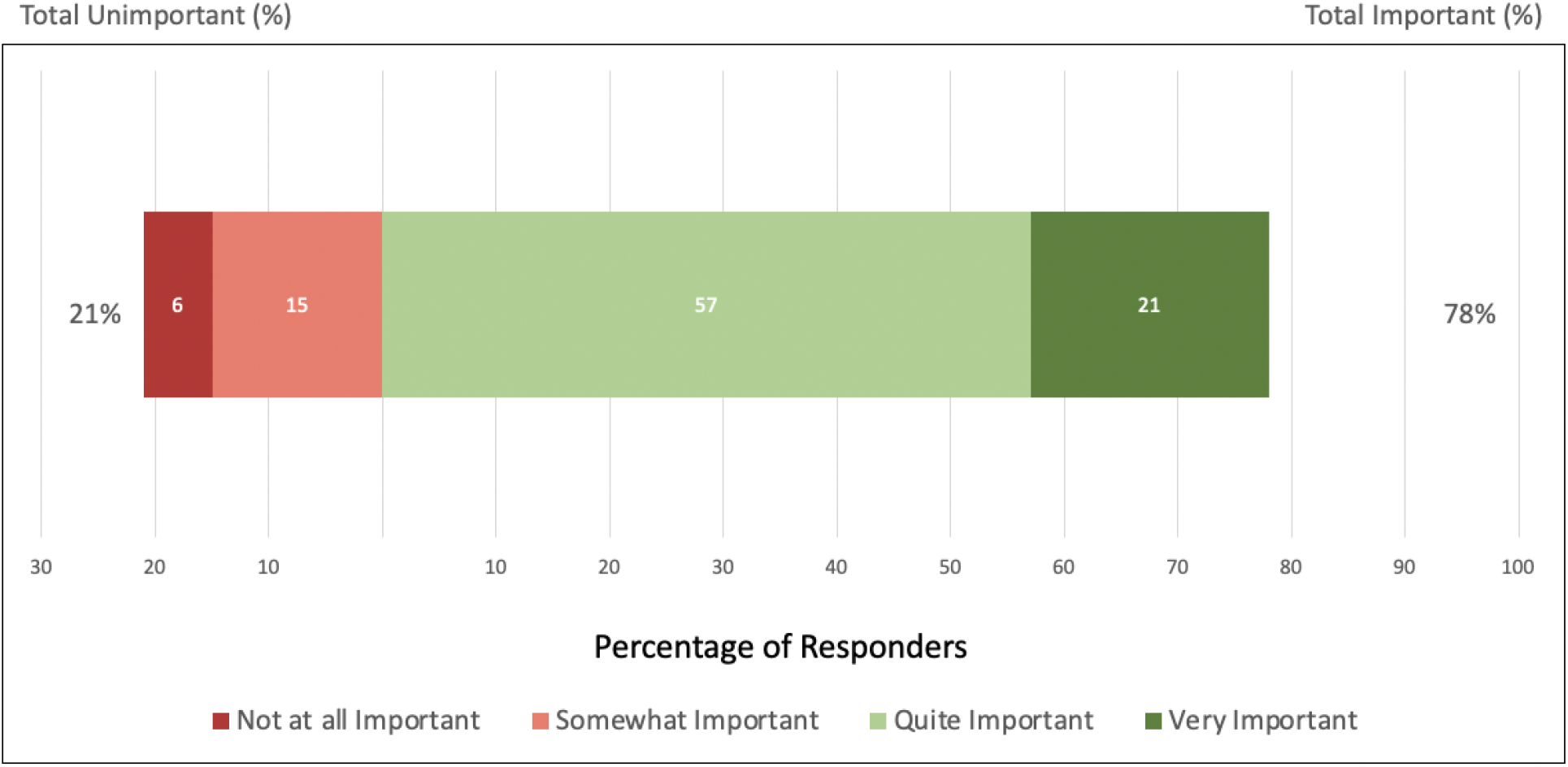
Responses about the importance of finding substitute non-medication therapies for maintenance treatment.

Figure 6 shows the responses to the vignette about an RCT for light and negative ion therapies for maintenance treatment (data in Supplemental Table S2). A total of 174 (78%) respondents endorsed that they would be “Very likely,” “Moderately likely,” or “Slightly likely” to volunteer for the study.

**Figure 6.**
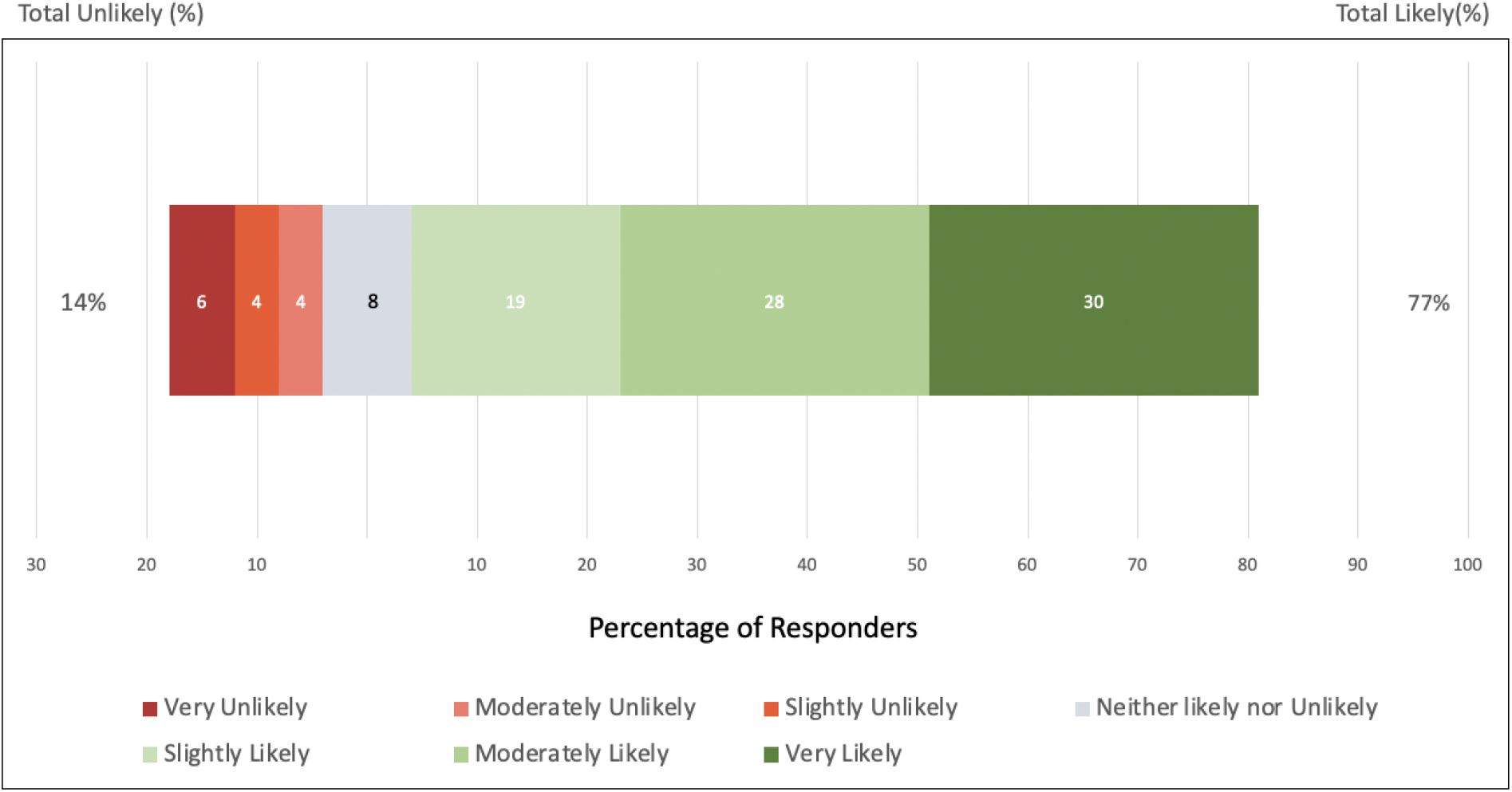
Responses about the likelihood to volunteer for the randomized study.

## DISCUSSION

This online survey examined attitudes about light therapy and negative ion therapy in participants who self-reported a diagnosis of depression. The demographic profile of participants (middle-aged, female, White/European ethnicity, university educated) mirrors those of most RCTs for depression treatments. This sample of participants was generally familiar with both treatments, although more participants had heard of light therapy than negative ion therapy (95% versus 62%, p<0.0001) and more had used light therapy (28% versus 16%, p<0.003). Similarly, the effectiveness of light therapy as similar to, or better than, antidepressants was endorsed by a larger percentage of the sample than for negative ion therapy (54% versus 37%, p<0.001), with more “unsure” about the effectiveness of negative ion therapy (34% versus 19%, p<0.001). These differences may be expected since light therapy has been widely reported in the media as an effective treatment for SAD and is endorsed by clinical guidelines for depression (e.g., Ravindran et al., 2016), whereas negative ion therapy is still largely limited to research studies.

Both treatments were regarded by respondents as easy to use, with no significant differences between the two. The reasons for wanting to use the treatments were similar across the two therapies, suggesting that they represent general attitudes about non-medication therapies versus medications rather than specifically for one therapy over the other. Having fewer side effects was the top reason for wanting to use both therapies. Reasons for not wanting to use light therapy and negative ion therapy were again similar for both, including apprehension about their effectiveness and about withdrawal effects from stopping antidepressants. Only 16% of the sample were concerned about the cost of each device or about side effects.

Finally, the sample felt strongly about the need for finding alternative non-medication treatments for relapse prevention of depression, with over three quarters of the sample (78%) finding this an important question to study. A similar percentage of participants (77%) was likely to volunteer for an RCT for maintenance treatment using one of these therapies.

This study had limitations, including a non-representative sample recruited by email and social media, which limits the participants to those with internet access. The diagnosis of depression was based on self-report, and it is unclear whether the treatments previously used were prescribed or supervised by health care professionals.

In conclusion, this online survey showed that people with depression had good familiarity with light therapy and negative ion therapy. Participants expressed positive attitudes towards their potential for maintenance treatment of depression, especially their ease of use. There was endorsement of the importance for finding alternative non-medication treatments to substitute for antidepressants for maintenance treatment, and the majority would likely participate in a randomized study. These findings suggest that it is important and feasible to conduct an RCT to investigate the efficacy and safety of these treatments for relapse prevention in patients with MDD.

## Data Availability

Data are available upon request.

## ACKNOWLEDGEMENTS

This study was funded by the Michael Smith Foundation for Health Research, award P2P18925. IL was funded by a Mach-Gaensslen Foundation of Canada Award from the UBC Faculty of Medicine Summer Student Research Program.

## Appendix 1. Selected questions and vignettes from online survey

### Light therapy is a treatment for depression that uses daily exposure to bright light from a light box deviceused at home. Light therapy usually has fewer side effects than antidepressant medications

***Q17*. Have you heard of light therapy for depression?**

◯ Yes, and I have used light therapy
◯ Yes
◯ No

***Q18. In your opinion, how effective is light therapy for depression treatment?***

◯ More effective than antidepressants
◯ As effective as antidepressants
◯ Less effective than antidepressants
◯ Not effective
◯ Unsure

### Negative ion therapy is a treatment for depression that uses daily exposure to negative ions from a negative ion device used at home. Negative ion therapy usually has fewer side effects than antidepressant medications

***Q19*. Have you heard of negative ion therapy for depression?**

◯ Yes, and I have used negative ion therapy
◯ Yes
◯ No

***Q20*. In your opinion, how effective is negative ion therapy for depression treatment?**

◯ More effective than antidepressants
◯ As effective as antidepressants
◯ Less effective than antidepressant
◯ Unsure

### Consider this scenario for the following question

Terry is diagnosed with major depressive disorder (clinical depression) by the doctor and is treated withan antidepressant medication. After taking the pills for about three months, Terry is feeling much better and no longer has depressive symptoms. Terry’s doctor recommends continuing the antidepressant for another 6-12 months. This “maintenance treatment” will help prevent the depression from returning.

***Q21*. In your opinion, how important is it to substitute an evidence-based <u>non-medication treatment</u> for antidepressants for maintenance treatment?**

◯ Very important
◯ Quite important
◯ Somewhat important
◯ Not at all important

### Consider this scenario for the following questions

Instead of taking medications for maintenance treatment, Terry uses light therapy. Terry buys a light box from the pharmacy for about $100 and uses it at home as prescribed. Terry sits in front of the light box for 30 minutes in the morning soon after waking up. Terry can read or eat breakfast while using the light box, but must be awake. Terry uses light therapy every weekday, but not on weekends.

***Q22*. How easy would it be for you to use light therapy as described?**

◯ Very easy
◯ Moderately easy
◯ Slightly easy
◯ Neither easy nor difficult
◯ Slightly difficult
◯ Moderately difficult
◯ Very difficult

### Consider this scenario for the following questions

Instead of taking medications for maintenance treatment, Terry uses negative ion therapy. Terry buys a negative ion device from the pharmacy for about $100 and uses it at home as prescribed. Terry sits in front of the negative ion device for 30 minutes in the morning soon after waking up. Terry can read or eatbreakfast while using the negative ion device, but must be awake. Terry uses negative ion therapy everyweekday, but not on weekends.

***Q25*. How easy would it be for you to use negative ion therapy as described?**

◯ Very easy
◯ Moderately easy
◯ Slightly easy
◯ Neither easy nor difficult
◯ Slightly difficult
◯ Moderately difficult
◯ Very difficult

### Consider this scenario for the following questions

You have been diagnosed with major depressive disorder (clinical depression) by your doctor and have been treated with an antidepressant medication. After taking the pills for about three months, you feel much better and you no longer have depressive symptoms. Your doctor tells you that you should have maintenance treatment for 6-12 months to help prevent the depression from returning. You are now asked to volunteer for a research study to find out whether light therapy or negative ion therapy is effective as a substitute for antidepressants for maintenance treatment. In this study, you will slowly stop taking the antidepressant medication and be randomly assigned (like the flip of a coin) to treatment with light therapy or negative ion therapy for up to 12 months. You will takehome the light device or negative ion device to use as described in the previous questions. However, half of the devices have been deactivated so that they are not effective. Therefore, you have a 1 in 2 chance of using an ineffective maintenance treatment. You will complete questionnaires online (on a computer or mobile device) once a week (which takes about 15 minutes) and will come to the research clinic for a visit once a month (which takes about 1 hour). These questionnaires and visits will check whether your depression symptoms are returning. If thedepression symptoms return, you will stop the study and be treated with usual medical care.

***Q29*. How likely would you be to volunteer for this study?**

◯ Very likely
◯ Moderately likely
◯ Slightly likely
◯ Neither likely nor unlikely
◯ Slightly unlikely
◯ Moderately unlikely
◯ Very unlikely

## SUPPLEMENTAL TABLES

**Table S1.** Attitudes about Treatment.

|  | Frequency | Percentage |
| --- | --- | --- |
| **Effectiveness of light therapy** |  |  |
| More effective than antidepressants | 28 | 13 |
| As effective as antidepressants | 91 | 41 |
| Less effective than antidepressants | 56 | 25 |
| Not effective | 3 | 1 |
| Unsure | 41 | 19 |
| **Effectiveness of negative ion therapy** |  |  |
| More effective than antidepressants | 13 | 6 |
| As effective as antidepressants | 68 | 31 |
| Less effective than antidepressants | 55 | 25 |
| Not effective | 9 | 4 |
| Unsure | 74 | 34 |
| **Ease of using light therapy** |  |  |
| Very easy | 41 | 19 |
| Moderately easy | 94 | 43 |
| Slightly easy | 58 | 26 |
| Neither easy nor difficult | 9 | 4 |
| Slightly difficult | 12 | 5 |
| Moderately difficult | 4 | 2 |
| Very difficult | 1 | 1 |
| **Ease of using negative ion therapy** |  |  |
| Very easy | 46 | 21 |
| Moderately easy | 72 | 33 |
| Slightly easy | 58 | 26 |
| Neither easy nor difficult | 20 | 9 |
| Slightly difficult | 13 | 6 |
| Moderately difficult | 4 | 2 |
| Very difficult | 2 | 1 |
| **Wanting to use light therapy instead of medications** |  |  |
| It is a non-medication treatment | 84 | 38 |
| It has fewer side effects | 111 | 50 |
| I don't like taking medications | 70 | 32 |
| I have side effects from medication | 57 | 26 |
| It seems like a natural treatment | 58 | 26 |
| **Wanting to use negative ion therapy instead of medications** |  |  |
| It is a non-medication treatment | 81 | 37 |
| It has fewer side effects | 86 | 39 |
| I don't like taking medications | 76 | 34 |
| I have side effects from medication | 67 | 30 |
| It seems like a natural treatment | 38 | 17 |
| **What would stop from using light therapy instead of medications** |  |  |
| I'm worried it won't work | 75 | 34 |
| I don't like the side effects | 36 | 16 |
| I don't have time to do light therapy | 55 | 25 |
| I can't afford to buy a light device | 35 | 16 |
| I'm worried about withdrawal effects from stopping the antidepressants | 53 | 24 |
| **What would stop from using negative ion therapy instead of medications** |  |  |
| I'm worried it won't work | 83 | 38 |
| I don't like the side effects | 37 | 17 |
| I don't have time to do light therapy | 56 | 25 |
| I can't afford to buy a light device | 35 | 16 |
| I'm worried about withdrawal effects from stopping the antidepressants | 58 | 26 |

**Table S2.** Attitudes about Study in Vignette.

|  | Frequency | Percentage |
| --- | --- | --- |
| **Importance to find substitute treatment** |  |  |
| Very important | 46 | 21 |
| Quite important | 125 | 57 |
| Somewhat important | 32 | 15 |
| Not at all important | 14 | 6 |
| **Likely to volunteer for this study** |  |  |
| Very likely | 66 | 30 |
| Moderately likely | 62 | 28 |
| Slightly likely | 42 | 19 |
| Neither likely nor unlikely | 18 | 8 |
| Slightly unlikely | 9 | 4 |
| Moderately unlikely | 8 | 4 |
| Very unlikely | 14 | 6 |

## REFERENCES

GBD 2017 Disease and Injury Incidence and Prevalence Collaborators. Global, regional, and national incidence, prevalence, and years lived with disability for 354 diseases and injuries for 195 countries and territories, 1990–2017: a systematic analysis for the Global Burden of Disease Study 2017. Lancet 2018; 392:1789–1858.

Goel N, Terman M, Terman J, et al. Controlled trial of bright light and negative air ions for chronic depression. Psychol Med 2005; 35:945–55.

Kato M, Hori H, Inoue T, et al. Discontinuation of antidepressants after remission with antidepressant medication in major depressive disorder: a systematic review and meta-analysis. Mol Psychiatry 2021; 26:118–133.

Kennedy SH, Lam RW, McIntyre RS, et al. CANMAT 2016 clinical guidelines for the management of adults with major depressive disorder. Section 3. Pharmacological treatments. Can J Psychiatry 2016; 61:540–560.

Lam RW, Levitt AJ, Levitan RD, et al. Efficacy of bright light treatment, fluoxetine, and the combination in patients with nonseasonal major depressive disorder: a randomized clinical trial. JAMA Psychiatry 2016; 73:56–63.

Li K, Wei Q, Li G, et al. Time to lack of persistence with pharmacological treatment among patients with current depressive episodes: a natural study with 1-year follow-up. Patient Prefer Adherence 2016; 10:2209–2215.

Olfson M, Marcus SC, Tedeschi M, et al. Continuity of antidepressant treatment for adults with depression in the United States. Am J Psychiatry 2006; 163:101–8.

Perera S, Eisen R, Bhatt M, et al. Light therapy for non-seasonal depression: systematic review and meta-analysis. BJPsych Open 2016; 2:116–126.

Perez V, Alexander DD, Bailey WH. Air ions and mood outcomes: a review and meta-analysis. BMC Psychiatry 2013; 13:29.

Ravindran AV, Balneaves L, Faulkner G, et al. Canadian Network for Mood and Anxiety Treatments (CANMAT) 2016 Clinical Guidelines for the Management of Adults with Major Depressive Disorder: Section 5. Complementary and Alternative Medicine Treatments. Can J Psychiatry 2016; 61:576–87.

Samples H, Mojtabai R. Antidepressant self-discontinuation: results from the collaborative psychiatric epidemiology surveys. Psychiatr Serv 2015; 66:455–62.

Sim K, Lau WK, Sim J, et al. Prevention of relapse and recurrence in adults with major depressive disorder: Systematic review and meta-analyses of controlled trials. Int J Neuropsychopharmacol 2016; 19:pyv076.

Tao L, Jiang R, Zhang K, et al. Light therapy in non-seasonal depression: An update meta-analysis. Psychiatry Res 2020; 291:113247.

Terman M, Terman JS. Light therapy for seasonal and nonseasonal depression: Efficacy, protocol, safety, and side effects. CNS Spectrums 2005; 10:647–663.

